# Multiple genome-wide association studies of type 2 diabetes implicate several genes are associated with diabetic retinopathy based on UK Biobank

**DOI:** 10.1101/2023.12.02.23299320

**Authors:** Tengda Cai, Qi Pan, Yiwen Tao, Charvi Nangia, Aravind Lathika Rajendrakumar, Tania Dottorini, Mainul Haque, Colin Palmer, Weihua Meng

## Abstract

**Purpose:** To identify the genetic variants associated with diabetic retinopathy in type 2 patients from the UK Biobank cohort (*n* = 17,015) and supporting replication cohorts GODARTS (*n* = 5,013), GOSHARE (*n* = 1,754), Caucasian Australians (*n* = 518), FinnGen (*n* = 206,664) and Chinese (n = 1,007).

**Methods:** Totally eleven genome-wide association studies were applied to search for significant genetic variants.

**Results:** We found 5 different loci associated with type 2 diabetic retinopathy in or nearest gene *EYA2*, *MPDZ*, *NTNG1*, *CTAGE14P* and *MREGP1*. In the primary GWAS, a significant SNP rs6066146 located in gene *EYA2* showed a *p* value of 4.21 × 10^−8^ and may play a role in the development of the disease, with “spleen” reaching a significant level produced by tissue expression analysis. Corresponding heritability of DR was estimated to be 26.73% by SumHer. Among five genes, we found that genes *EYA2*, *MPDZ*, *NTNG1* had genetic interactions and may affect the complex development of retinal blood vessels.

**Conclusion:** Diabetic retinopathy is a complication of diabetes that affects the eyes. It is highly likely to occur when high blood sugar damages the retinal blood vessels. There is limited awareness regarding the pathogenesis of DR. Our study identified multiple loci associated with diabetic retinopathy, which may lead to personalized treatments to reduce the burden of the disease.

## Introduction

Diabetic retinopathy (DR), a disease influenced by multiple genes, stands as one of the most frequent ocular disorders in individuals suffering from diabetes. Around 35% of the global diabetic population is affected by DR, which is a leading cause of blindness in the workforce-aged demographic, despite the implementation of screening and treatments (Yau et al. 2012; Bunce et al. 2010). If left untreated, half of all patients diagnosed with proliferative diabetic retinopathy (PDR) are projected to become blind within five years post-diagnosis (Williams et al. 2004). As the global population ages and diabetes becomes more prevalent, it is anticipated that instances of DR will rise.

In diabetic patients, diabetic retinopathy is a common microvascular complication and is associated with an increased risk of life-threatening systemic vascular complications (Su et al. 2023). Hyperglycemia is known to lead to changes in the retina, causing vascular permeability, inflammation, fluid leakage and ischemia, and that certain growth factors may play a key role in diabetic microvascular complications (Pathak et al. 2012). Hyperglycemia and dyslipidemia are thought to disturb the homeostasis of the retina by inducing inflammatory responses in retinal tissue, including oxidative stress (Heesterbeek 2012). Epidemiologic studies have suggested multiple risk factors associated with the development and progression of diabetic retinopathy from longitudinal studies, including higher blood glucose, higher blood pressure, nonsmoking, male gender, higher HbA1c, longer duration of diabetes, lower BMI, and higher blood urea concentration (Meng et al. 2018). Moreover, many risk factors for DR have been reported recently, including hypertension, hyperlipidemia, poor blood glucose control, and albuminuria, although the underlying genetic mechanisms responsible for DR have not been identified (Looker et al. 2003; Leske et al. 2005; Cikamatana et al. 2007; Singh et al. 2008). Tentative risk factors included poor glycaemic control and a longer duration of diabetes (Stephen et al. 2018).

Studies have shown that patients with 20 years of diabetes, almost all type 1 diabetes (T1D) patients and 58% of type 2 diabetes (T2D) patients have signs of diabetic retinopathy (Hletala et al. 2010). Both twin studies and family studies have substantiated that the susceptibility to DR is genetically inherited in patients with type 1 and type 2 diabetes (Monti et al. 2007; Cho et al. 2014). In individuals, the retinal disorder progresses from non-proliferative diabetes to vision-threatening proliferative diabetes, which is characterized by the growth of abnormal new blood vessels in the retina (Liu et al. 2019). The new blood vessels and the ensuing contraction of fibrous tissue can deform the retina, leading to traction retinal detachment and severe, often irreversible, vision loss (Liu et al. 2019). Notably, the genetic influence is more pronounced in the severe forms of DR, with the heritability factor rising from 18% in non-PDR stages as sibling models, to a substantial 50% in PDR cases (Looker et al. 2012; Hallman et al. 2005; Hietala et al. 2008). Studies have shown that proliferative DR has a 52% heritability rate in families, suggesting that family genetic factors are also associated with DR. (Looker et al. 2007; Arar et al. 2008).

Lately, various genome-wide association studies (GWASs) have suggested numerous potential genetic loci linked to DR, yet none of them have attained genome-wide significance or been confirmed by replication studies (Fu et al. 2010; Grassi et al. 2011; Awata et al. 2014; Burdon et al. 2015; Meng et al. 2018; Pollack et al. 2019). Furthermore, none of the linkage analyses, candidate gene association studies, and genome-wide association studies have identified DR risk loci that can be consistently reproduced. Insufficient grasp of DR’s genetic aspects impedes the discovery of new biological routes for intervention, consequently escalating the healthcare expenses related to DR.

To better understand the genetic mechanisms related to diabetic retinopathy with type 2 mellitus, we carried out multiple GWASs using the UK Biobank (UKB) cohort. So far, this is a novel GWAS study conducted on the UKB cohort specifically for diabetic retinopathy. This project is designed to explore the genetic loci associated with DR based on the UK Biobank cohort and will give new ideas to those researchers studying DR.

## Materials and Methods

### Information of Cohorts and Patients

The UK Biobank serves as a large-scale biomedical database and research resource, offering comprehensive genetic and phenotypic data on roughly half a million UK participants aged between 40 and 69 years across England, Scotland, and Wales. For additional details regarding the UK Biobank cohort, refer to the website www.ukbiobank.ac.uk. This study was conducted in accordance with the Tenets of the Declaration of Helsinki. Ethical approval for this research was appropriately secured from the National Health Service National Research Ethics Service (reference 11/NW/0382). Current studies were conducted based on UKB cohorts.

In this research, the gene data came from the UK Biobank cohort. They used a standardized process for DNA extraction and quality control (QC), in which detailed methods can be found at https://biobank.ctsu.ox.ac.uk/crystal/ukb/docs/genotyping_sample_workflow.pdf. Type 2 diabetic patients were identified as individuals who responded with “No” to a specific query about “Started insulin within one year diagnosis of diabetes” in UK Biobank questionnaire (Refer to UK Biobank Questionnaire field ID: 2986). That is, individuals classified as type 2 diabetic patients are those who reported not commencing insulin therapy within a year of diagnosis. Patients with DR are diagnosed and collective according to the ICD 10 - WHO International Classification of Diseases (Refer to UK Biobank Questionnaire field ID: 41202). Moreover, we focused our study on participants of White British (Refer to UK Biobank Questionnaire field ID: 21000).

Upon extracting data from the UKB cohort, the numbers and clinical characteristics of cases and controls would be shown in Table S1 with significant GWASs. The case-control definition compared any DR to no DR. Cases were identified by participants who had T2D as well as DR while control subjects were distinguished by participants who had T2D but not DR.

During the replication phase, it is challenging to obtain summary statistics of DR patients with type 2 mellitus, as most provide only small datasets of the top loci (e.g., p < 1×10^−5^) and limited researchs. We attempted to expand our collection to different datasets for DR research which may not match exactly in definition but favored exploring the replication of loci. we matched the significant loci of four GWAS studies on diabetic retinopathy of mainly type 2 patients with our results. The studies are using the following cohorts: 1. GoDARTS (evolved from the Diabetes Audit and Research in Tayside Scotland setup in 1996 jointly by the University of Dundee etc.) and GoSHARE (GODRTS recruitment began in 1998 intending to obtain genotypic information on individuals with diabetes) (Rajendrakumar unpublished). 2. Caucasian Australians (funded by the National Health and Medical Research Council of Australia (GNT 595918) (Graham et al. 2018). 3. FinnGen cohorts (based on a public-private partnership between Finnish universities, biobanks, hospital districts, and several international pharmaceutical companies) (FinnGen. GWAS summary results: diabetic retinopathy). 4. Chinese cohorts (from Taiwan-US Diabetic Retinopathy study) (Sheu et al. 2013). More information about these cohorts and GWASs was shown in Table 1.

**Table 1.** Information of four GWASs (five cohorts) in replication phase.

| Cohorts | Studied Trait | Background Disease | Definition of Cases and Controls | Discovery Sample Size (N) |  |  |
| --- | --- | --- | --- | --- | --- | --- |
|  |  |  |  | Cases | Controls | Total |
| GoDARTS | DR | Primarily Type 2 Mellitus,<br>Few Type 1 Mellitus | Case: samples with any DR in T2D or T1D<br>Control: samples without DR in T2D or T1D | 1,791 | 3,222 | 5,013 |
| GoSHARE | DR | Primarily Type 2 Mellitus,<br>Few Type 1 Mellitus | Case: samples with any DR in T2D or T1D<br>Control: samples without DR in T2D or T1D | 527 | 1,227 | 1,754 |
| Caucasian Australians | PDR | Type 2 Mellitus | Case: samples with PDR in T2D<br>Control: samples without DR in T2D | 176 | 435 | 518 |
| FinnGen | DR | Type 1 & 2 Mellitus | Case: samples with DR<br>Control: samples without DR | 3,646 | 203,018 | 206,664 |
| Chinese | PDR | Type 2 Mellitus | Case: samples with PDR in T2D<br>Control: samples without DR in T2D | 437 | 570 | 1,007 |

### Definitions of multiple GWASs

When performing primary GWAS, the tool we used is genome-wide association methods based on generalized linear mixed model association tool (fastGWA-GLMM) (Yang et al. 2021), which in model covariates were age, sex, bmi, duration of diabetes (DoD), and top 8 genetic principal components. Based on the model in primary GWAS, there were two further GWASs generated from models adding other important covariates HbA1c and urea respectively as they have a high likelihood of impact in DR according to epidemiological studies (Roychoudhury et al. 2021; Meng et al. 2018). In most general sense, “covariates” typically refer to X variables that are included in a model to refine the precision of estimated treatment effects. In observational study designs, covariates might be added to a model to 1) to enhance predictive power, 2) to investigate specific conditional effects, or 3) to control for confounding (Walker 2018). Overall, the inclusion of covariates HbA1c and Urea respectively aims to increase the statistical power and validity of the model, ensuring the reliability of the research findings.

Considering the impact of data distribution in specific covariates, there are 8 secondary GWASs generated from different datasets. As we are interested in the datasets that have high age, HbA1c, Urea, and prolonged DoD which may help us to find out significant genetic loci with DR, datasets with age, DoD, HbA1c and Urea larger than corresponding Median (Q2) & Upper quartile (Q3) are mined out to perform GWAS respectively since the magnitude of these covariates is not clearly delineated in terms of the stages. Totally 11 different GWASs (3 GWASs and 8 Secondary GWASs) designed were shown in Table 2. The first row of the table represents three basic GWASs, where GWAS(+HbA1c) and GWAS(+Urea) indicate the addition of covariates HbA1c and Urea, respectively, to the linear model of the Primary GWAS. Second row shows secondary GWASs studies conducted using sub-datasets based on the model of the Primary GWASs. For example, in DoD>Q2, it means that the chosen sub-dataset is where the variable (duration of diabetes) is greater than its own median, based on the model used in the Primary GWAS.

**Table 2.** *Upper part* shows 3 GWASs using different models in fastGWA-GLMM. *Lower part* totally shows 8 secondary GWASs based on different dataset requirements in each grey part.

| GWASs | GWAS(+HbA1c) | Primary GWAS | GWAS(+Urea) |
| --- | --- | --- | --- |
| Secondary GWASs | HbA1c > Q2 | Age > Q2, Age > Q3 | Urea > Q2 |
|  | HbA1c > Q3 | DoD > Q2, DoD > Q3 | Urea > Q3 |

### Statistical analysis

Genome-Wide Complex Trait Analysis (GCTA v1.93.3), a package used in this research under Linux, is designed to estimate the proportion of phenotypic variation explained by all genome-wide SNPs for complex traits (refer to https://yanglab.westlake.edu.cn/software/gcta/#Overview). Our study performed fastGWA-GLMM for binary traits in UK Biobank data scale. In QC step, SNPs with imputation INFO scores < 0.3, minor allele frequency < 0.5% were removed by using GCTA, as well as SNPs that failed Hardy-Weinberg < 0.001 and samples with missing call rates < 0.1 would be filter out by using Plink v1.90. SNPs situated on the X and Y chromosomes, as well as mitochondrial SNPs, were eliminated from consideration. Gender difference between cases and controls was compared using chi-square testing, whereas age and BMI were compared by conducting independent t-testing with R v4.0.3. SNPs were considered reaching genome-wide association significance if they had a *p* value less than 5×10^−8^. Moreover, heritability was calculated by utilizing “BLD-LDAK Model” in SumHer (Speed & Balding 2018; Speed et al. 2020).

### GWAS analysis by FUMA and LocusZoom

GRCh37 genome assembly was followed for SNP annotation. SNP functional annotations, Manhattan plot and Q-Q plot were applied and produced by the FUMA v1.5.6, a web platform for annotating, prioritizing, visualizing and interpreting GWAS results (Watanabe et al. 2017).

Regional visualization was provided by LocusZoom (http://locuszoom.org/). The gene analysis, gene-set analysis and tissue expression analysis were performed with MAGMA v1.08, which was integrated in FUMA. In the gene analysis, the summary statistics of SNPs are consolidated to the level of entire genes, evaluating the combined association of all SNPs in that gene with the phenotype. Gene-set analysis groups individual genes based on shared biological, functional, or other traits. This helps in understanding the role of particular biological pathways or cell functions in the genetic origins of a phenotype. To test the relationship between highly expressed genes in a specific tissue and genetic associations with DR, tissue expression analysis, based on GTEx (https://www.gtexportal.org/home/), was performed separately for 30 general tissue types and 53 specific tissue types.

## Results

### Primary GWAS results

Through data extraction from questionnaire and quality control on gene data, we identified 1,824 cases and 15,164 controls. Clinical characteristics of the case and control groups were compiled in Table S1, Sheet 1. Age, sex, body mass index (BMI), duration of diabetes, HbA1c and Urea were all found to be significantly different (*p* < 0.005) between cases and controls. There was a cluster appearing of top SNPs in the Manhattan plot, where a locus reaches significant level (Fig. 1). Corresponding Q-Q plot is shown in Figure S1. The top SNP in this region was rs6066146 in the *EYA2* gene with a *p* value of 4.21×10^−8^ in Chromosome 20. Following suggestive SNPs are rs117659116 in chromosome 12 with a *p* value of 5.77×10^−8^ in gene *MREGP1*, rs17277029 in chromosome 4 with a *p* value of 6.01×10^−7^ in gene *FBXW7* and rs112773829 in chromosome 2 with a *p* value of 8.95×10^−7^ in gene *RNU6-546P*. Altogether 11,169,041 imputed SNPs passed from routine quality control checking and imputation INFO quality score larger than 0.3. The heritability of DR was estimated to be 26.73% in this type 2 diabetic population based on the combination of the Baseline LD model (Finucane et al. 2015; Gazal et al. 2019) and the LDAK model (Speed et al. 2020) by SumHer.

**Fig. 1.**
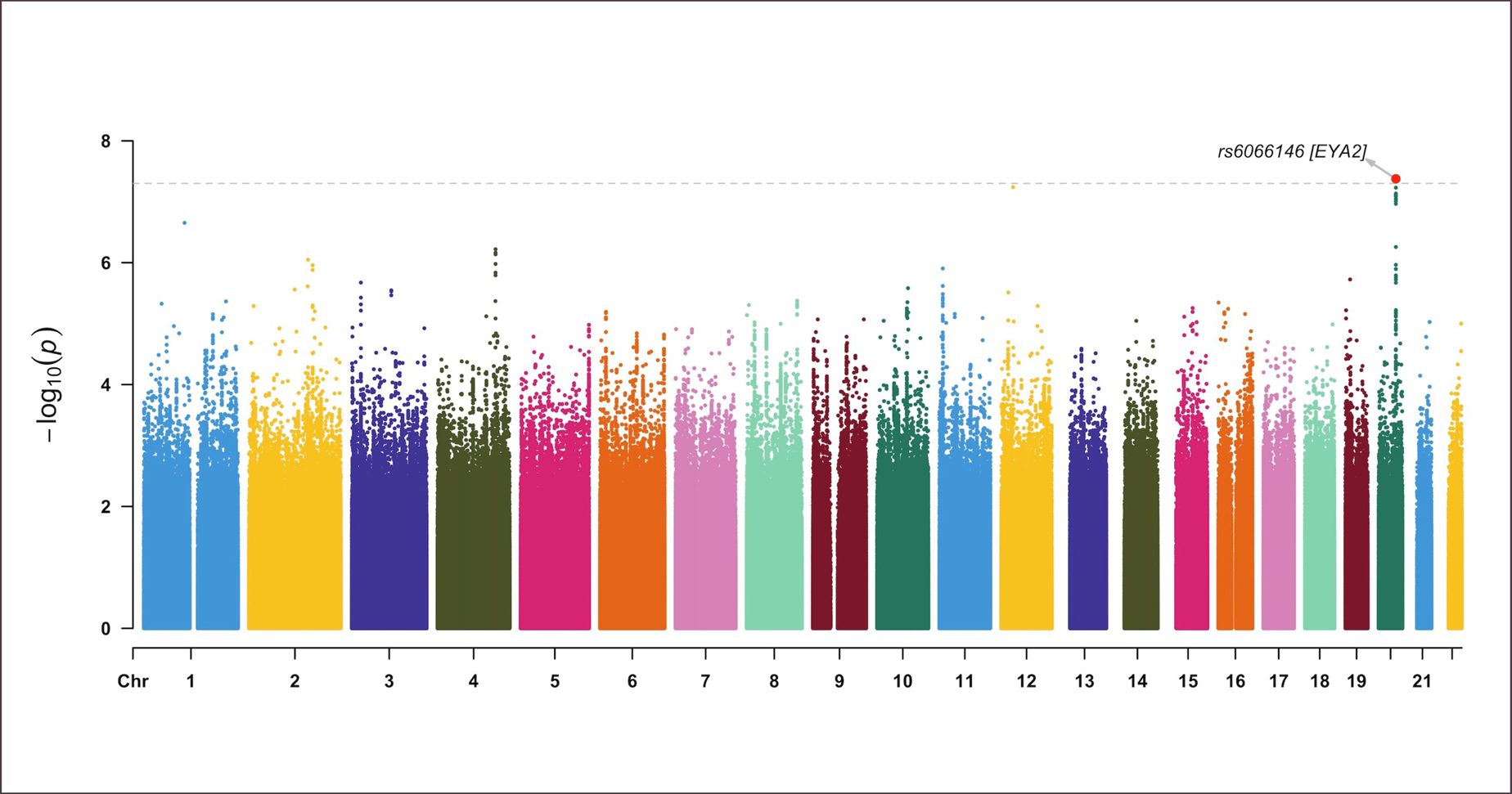
The Manhattan plot of the primary GWAS analysis on diabetic retinopathy in type 2 patients (N = 16,988) The dashed grey line indicates the cut-off *p* value of 5×10^−8^

### Gene, gene-set and tissue expression analysis by FUMA

The parameters in the FUMA platform are set as follows. N = 16988, exMHC = 1, ensembl = v102, genetype = protein_coding, leadP = 5e-8, gwasP = 0.05, r2 = 0.6, 2ndr2 = 0.1, refpanel = UKB/release2b, pop = WBrits_10k, MAF = 0, mergeDist = 250, magma = 1, magma_window = 42, magma_exp = GTEx/v8/gtex_v8_ts_avg_log2TPM, GTEx/v8/gtex_v8_ts_general_avg_log2TPM, posMap = 1, posMapWindowSize = 10.

In gene analysis, all the SNPs that are located within genes were mapped to 19,238 protein-coding genes. No gene demonstrated the strongest association, see Figure S2.

In the gene-set analysis, a total of 10,678 gene sets were tested. There is no gene set achieve statistically significant association (*p* < 0.05/10,678 = 4.68×10^−6^). The top first gene set are GOBP_NEGATIVE_REGULATION_OF_CELL_JUNCTION_ASSEMBLY demonstrated *p* value of 1.39×10^−5^. The top ten gene sets from this analysis are shown in Table S2.

In the tissue expression analysis, tissue “Spleen” demonstrated statistically significant associations in the expression analysis of 30 general tissue types from multiple organs and the 53 specific tissue types within some of these organs. See Fig. 2.

**Fig. 2.**
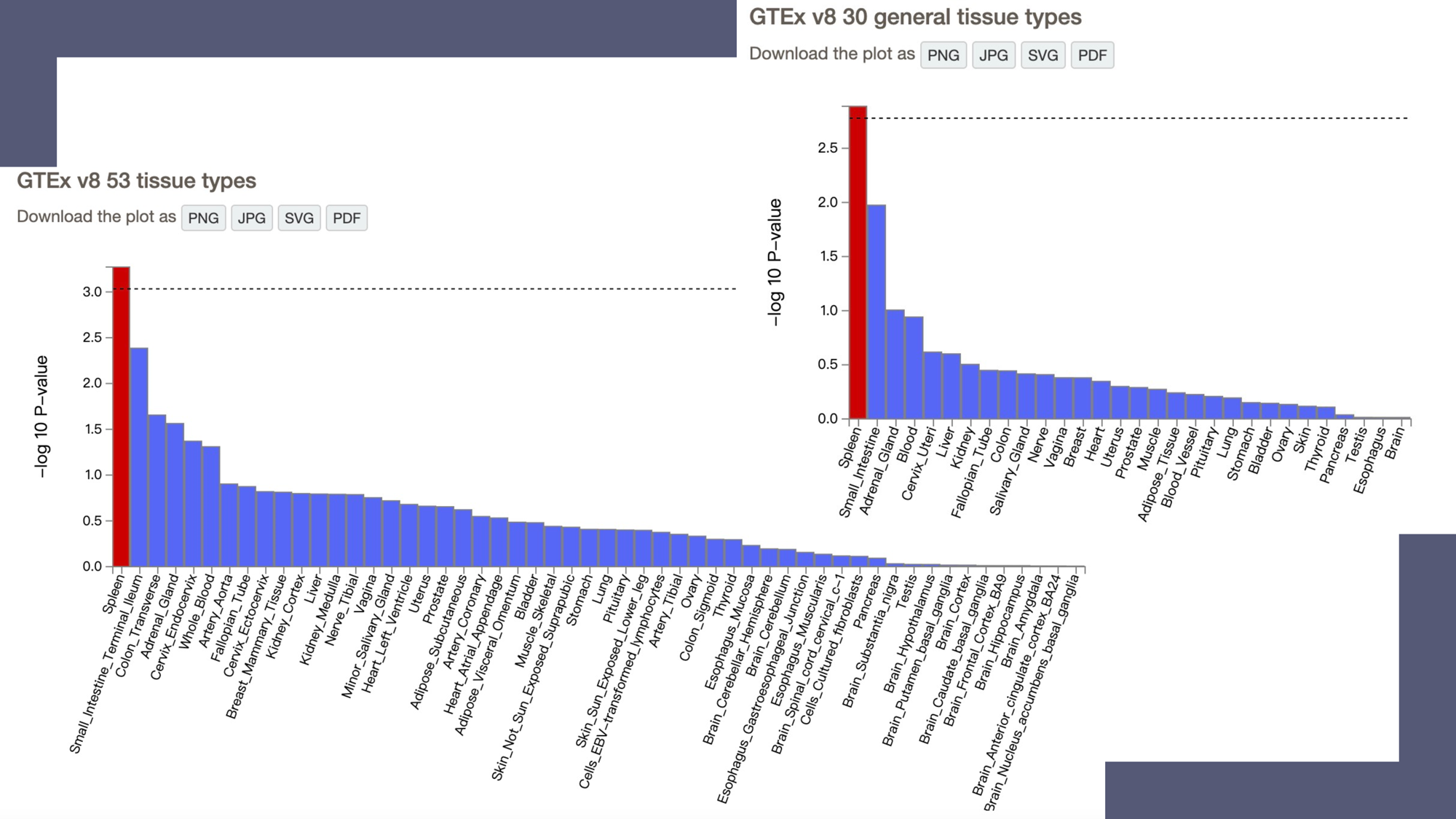
Tissue expression results on 30 general tissue types and 53 specific tissue types by GTEx in the FUMA. The dashed line shows the cut-off *p* value for significance with Bonferroni adjustment for multiple hypothesis testing

### Additional GWASs results

All GWASs which had significant results are concluded in Table 3. Fig. 3 shows circular Manhattan plot for GWAS(+HbA1c), GWAS(+Urea), primary GWAS(DoD>Q2), GWAS(HbA1c>Q3). Gene *EYA2* was shown to be a significant gene in several experiments. A Marker of 1: 107,782,413 with the nearest gene *NTNG1* is also shown to be a significant gene in several GWASs while there is a SNP cluster in both gene *CTAGE14P* and *MPDZ* respectively. All regional plots for 4 loci are shown in Figure S3, while Figure S4 shows the Q-Q plot of GWAS(+HbA1c), GWAS(+Urea), primary GWAS(DoD>Q2) and GWAS(HbA1c>Q3) respectively.

**Fig. 3.**
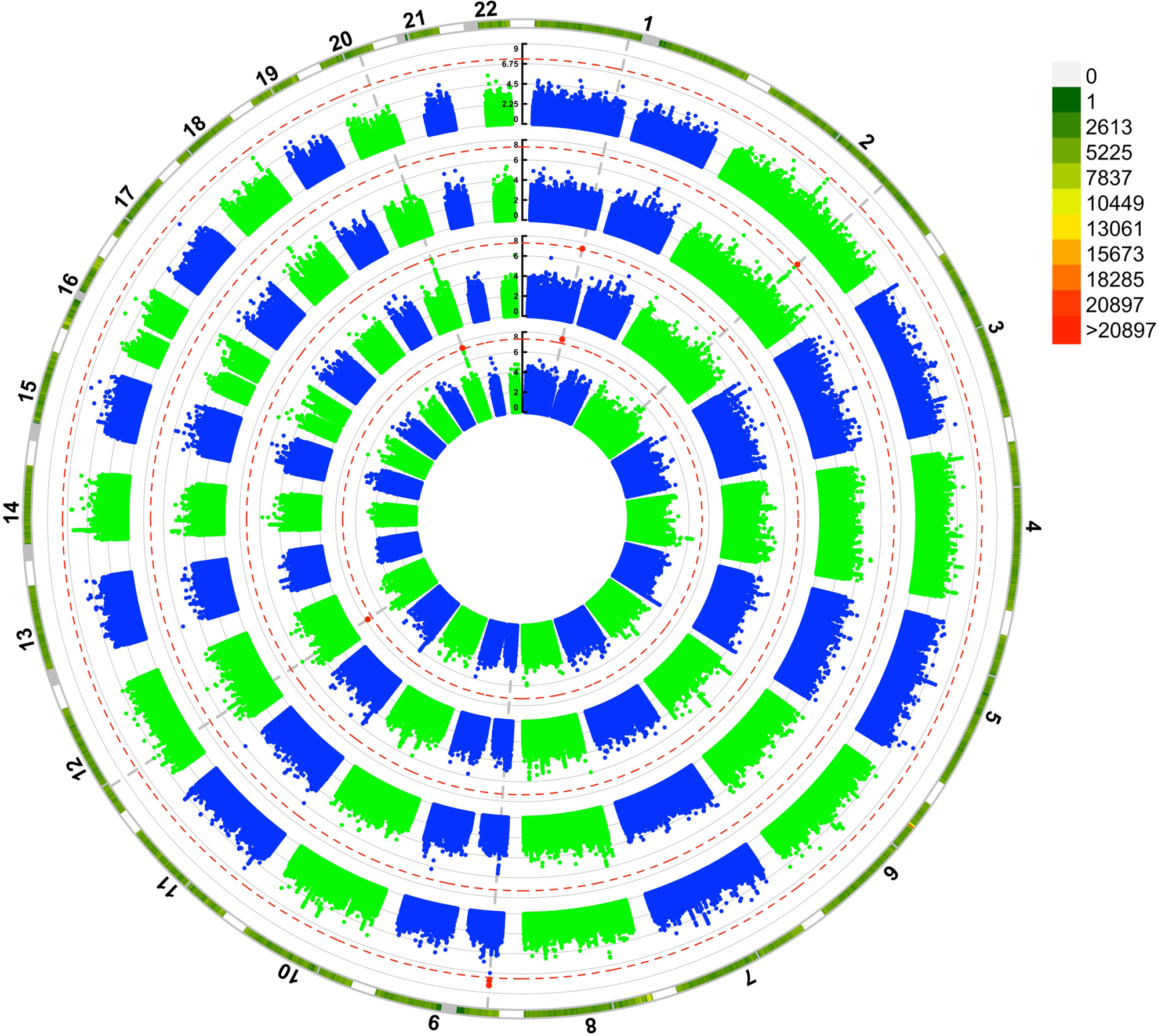
The Circular Manhattan plots of additional GWASs analysis on diabetic retinopathy in type 2 patients

**Table 3.** Significant and suggestive loci of all GWASs using UK Biobank.

| GWASs | Marker | Chromosome | Position | Nearest gene | p value |
| --- | --- | --- | --- | --- | --- |
| Primary GWAS | rs6066146 | 20 | 45,599,553 | *EYA2* | 4.21 x10<sup>-8</sup> * |
|  | rs117659116 | 12 | 31,418,552 | *MREGP1* | 5.77 x10<sup>-8</sup> |
|  | 1: 107,782,413 | 1 | 107,782,413 | *NTNG1* | 2.22 x10<sup>-7</sup> |
|  | rs17277029 | 4 | 153,428,649 | *FBXW7* | 6.01 x10<sup>-7</sup> |
|  | rs112773829 | 2 | 156,268,365 | *RNU6-546P* | 8.95 x10<sup>-7</sup> |
| GWAS (+HbA1c) | rs117659116 | 12 | 31,418,552 | *MREGP1* | 1.75 x10<sup>-8</sup> * |
|  | 1: 107,782,413 | 1 | 107,782,413 | *NTNG1* | 1.96 x10<sup>-8</sup> * |
|  | rs6066146 | 20 | 45,599,553 | *EYA2* | 3.77 x10<sup>-8</sup> * |
|  | rs72721645 | 4 | 153,494,209 | *AC023424.2* | 3.78 x10<sup>-7</sup> |
|  | rs552254054 | 16 | 549,951 | *RAB11FIP3* | 8.99 x10<sup>-7</sup> |
|  | 11: 9,199,268 | 11 | 9,199,268 | *DENND5A* | 9.66 x10<sup>-7</sup> |
| GWAS (+Urea) | 1: 107,782,413 | 1 | 107,782,413 | *NTNG1* | 4.08 x10<sup>-8</sup> * |
|  | rs71183224 | 20 | 45,603,386 | *EYA2* | 5.96 x10<sup>-8</sup> |
|  | rs6435333 | 2 | 155,560,333 | *KCNJ3* | 4.04 x10<sup>-7</sup> |
|  | rs34759185 | 1 | 217,293,468 | *ESRRG* | 6.28 x10<sup>-7</sup> |
|  | rs552254054 | 16 | 549,951 | *RAB11FIP3* | 7.17 x10<sup>-7</sup> |
|  | rs142927559 | 6 | 97,386,148 | *KLHL32* | 7.21 x10<sup>-7</sup> |
|  | rs137934083 | 22 | 49,479,088 | *AL078622.1* | 8.02 x10<sup>-7</sup> |
| Primary GWAS (DoD > Q2) | rs13013265 | 2 | 168,556,935 | *CTAGE14P* | 2.69 x10<sup>-8</sup> * |
|  | rs79202341 | 9 | 115,618,322 | *SNX30* | 5.18 x10<sup>-7</sup> |
|  | rs147264322 | 2 | 78,314,433 | *AC012494.1* | 8.09 x10<sup>-7</sup> |
|  | rs190386742 | 2 | 170,057,766 | *LRP2* | 9.08 x10<sup>-7</sup> |
| GWAS (HbA1c > Q3) | rs62532872 | 9 | 13,226,669 | *MPDZ* | 6.90 x10<sup>-9</sup> * |
|  | rs17273542 | 9 | 13,224,491 | *MPDZ* | 2.32 x10<sup>-8</sup> * |
|  | rs186759284 | 2 | 116,694,780 | *AC016721.1* | 1.19 x10<sup>-7</sup> |
|  | rs550645235 | 4 | 26,779,514 | *TBC1D19* | 1.63 x10<sup>-7</sup> |
|  | rs147617022 | 5 | 164,192,487 | *AC109466.1* | 2.89 x10<sup>-7</sup> |
|  | 17: 57,849,382 | 17 | 57,849,382 | *VMP1* | 3.94 x10<sup>-7</sup> |
|  | rs145145808 | 2 | 116,905,368 | *AC016721.1* | 4.23 x10<sup>-7</sup> |
|  | rs189997796 | 14 | 58,510,393 | *ARMH4* | 4.70 x10<sup>-7</sup> |
|  | rs117849333 | 18 | 70,439,751 | *NETO1* | 5.51 x10<sup>-7</sup> |
|  | rs77360543 | 15 | 76,160,587 | *UBE2Q2* | 5.62 x10<sup>-7</sup> |
|  | rs548908893 | 2 | 69,970,492 | *ANXA4* | 7.56 x10<sup>-7</sup> |

\* indicates significant loci reach significant p value 5x10<sup>-8</sup> , while others are suggestive loci.

### Replication Stage

In the GoDARTS and GoSHARE cohorts, rs6066146 was not significantly replicated, while rs17273542 had a p-value consistently less than 0.1. In the Caucasian Australians cohort, no loci were significantly replicated. In the FinnGen cohort’s study on VII Diseases of the eye and adnexa (H7_), both rs6066146 and rs71183224 had p-values less than 0.01, indicating suggestive replication. In Chinese cohorts, rs6066146 showed p value less than 0.1. Detailed p values of loci in these studies are shown in the Table S3.

## Discussion

In these GWAS studies of type 2 diabetic retinopathy using the UKB dataset, our research group identified noteworthy genetic variants at five different loci. These findings highlight the complex genetic basis associated with DR. Notably, in this study we used the UKB’s generic questionnaire as a valuable screening tool, providing researchers with the advantage of mitigating the potential problem of reduced study power due to heterogeneity. Results showed that in primary GWAS on diabetic retinopathy, we identified variants in *EYA2* have reached genome-wide significance (*p* < 5×10^−8^). In addition, other GWASs further revealed that variants in or near gene *MREGP1*, *MPDZ*, *NTNG1* and *CTAGE14P* were identified to be involved in the development of DR.

Leading locus rs6066146 in gene *EYA2* in chromosome 20 for DR was identified from primary GWAS. The *EYA2* gene, which stands for “Eyes Absent Homolog 2”, encodes a member of the eyes absent family of proteins, which encodes proteins that may be post-translationally modified and may play a role in eye development (Wang et al. 2016). *EYA2* is involved in DNA repair and has, besides type 2 diabetes, also been reported to associate with triglyceride levels and waist-hip ratio (Jonsson et al. 2021). Zhang et al. had found that *EYA2* is also expressed in other sensory and developmental systems (eye) that have not been reported to date. The research showed that in the retina, X-gal staining of *EYA2^lacZ^*at E12.5 and E14.5 showed β-Gal activity in *EYA2^lacZ/+^* and *EYA2^lacZ/lacZ^*, as well as β-Gal activity was also observed in the sclera, extraocular muscles, and nerve fibers within the optic stalk, which will develop into optic nerve (Zhang et al. 2021). Studies above supported a possible role of *EYA2* in retinal neuronal differentiation. To expand on that, the four human *EYA* paralogs (*EYA1-4*) are initially elucidated within the framework of Drosophila eye development as the retinal determination gene network (Bonini et al. 1993). Research findings indicate that the pharmacological suppression of *EYA* protein tyrosine phosphatase (PTP) activity leads to a slowdown in vascular growth within the postnatal mouse retina (Roychoudhury et al. 2021). An experiment in a mouse model of oxygen-induced retinopathy demonstrated that *EYA* inhibitors reduced pathological neovascularisation in an in vivo model for proliferative retinopathy (Hegde & OH 2017). Observations above sheds light on the potential impact of *EYA2* on human retinal vasculature. Additionally, the *EYA* tyrosine phosphatase activity plays a crucial role in deciding between repair and apoptosis following DNA double-stranded breaks, which activity might promote hypoxia-induced angiogenesis by dephosphorylating H2AX, which implies that *EYA* is likely involved in both developmental and disease-related retinal angiogenesis (Wang et al. 2016). In summary, many studies showcase the pro-angiogenic nature of *EYA* tyrosine phosphatase activity (Roychoudhury et al. 2021; Wang et al. 2016; Tadjuidje et al. 2012). Both Benzbromarone and Benzarone emerge as promising contenders for repurposing as potential drugs to address cancer metastasis, tumor-driven angiogenesis, and vascular disorders (Tadjuidje et al. 2012).

Tissue expression analysis facilitated by FUMA indicated an association between ‘Spleen’ and gene *EYA2*, where *EYA PTP* activity had been identified (Pandey et al. 2013; Tadjuidje et al. 2012), which is associated with angiogenesis (Wang et al. 2018) and proliferative retinopathy (Wang et al. 2016) and have been validated in animal disease models. Recent studies had elaborated that microglial spleen tyrosine kinase might serve as a potential therapeutic target for diabetic retinopathy (Liu et al. 2021; Su et al. 2020), which means that some physiological change in the spleen to some degree might contribute to the development of DR. Another experimental study showed that in diabetic patients, circulating pro-inflammatory monocytes of bone marrow origin were increased, whereas reparative circulating angiogenic cells were trapped in the bone marrow and spleen and had an impaired ability to be released into the circulatory system, leading to inadequate retinal vascular repair, that is, developing diabetic retinopathy (Chakravarthy et al. 2016). Moreover, in the type 2 diabetic individuals, the heritability of DR was calculated to be 26.73%. It is noteworthy that this heritability estimation falls within the range of heritability reported by most studies (Simó-Servat et al. 2013; Looker et al. 2012; Hietala et al. 2008; Hallman et al. 2005).This assessment does not encompass the contribution of gene-environment interactions, etc., implying that the genuine heritability of this trait might be higher. We have posited that DR exhibits hereditary characteristics in this GWAS, underscoring the need for additional genetic investigations.

Significant loci found in other GWASs were as follows. Variant rs117659116 in gene *MREGP1* in chromosome 12 and variants rs62532872, rs17273542 in *MPDZ* gene in chromosome 9. *MREGP1* have been pointed out twice in the GWAS studies, but there were no others supporting except that the gene was found out associated with type 2 diabetes mellitus, with rs117659116 could not found any information about DR. In addition, we have found two significant loci, which are the Marker 1: 107,782,413 in chromosome 1 near gene *NTNG1*, and the locus rs13013265 near gene *CTAGE14P* in chromosome 2. Gene *NTNG1* (Netrin G1), identified by GWAS(+HbA1c) and GWAS(+Urea), was a protein-coding gene. A study shows that the expression of *NTNG1* was high in the retina and kidney in GTEx (https://www.gtexportal.org/home), and in the brain and kidney in HPA-RNA (Ke et al. 2023). Abnormal methylation-modified differentially expressed genes of *NTNG1* may be involved in inflammatory response and vascular remodeling in the process of Coronary artery ectasia (Yang et al. 2023), which may likely influence the retinal vasculature. Gene *CTAGE14P* was identified from GWAS(DoD>Q2), which was done to explore whether the case and control group would show significant differences in a group of people who have had diabetes for more than 5 years. Though gene *CTAGE14P* has been found to be associated with visual perception in previous studies (Zhu et al. 2020), there is not enough evidence that the gene is associated with DR. DR patients were somewhat insensitive to age according to the secondary GWASs in age. For all secondary GWASs(+Urea), we did not find any significant loci to support the involvement of high urea in the development of DR.

The *MPDZ* gene was obtained from a study GWAS(HbA1c>Q3), which aimed to explore whether there was a significant difference between the case and control group in DR patients with high levels of HbA1c. *MPDZ* (Multiple PDZ Domain Crumbs Cell Polarity Complex Component) is a protein-coding gene. Diseases associated with *MPDZ* include hydrocephalus, congenital, 2, with or without brain or eye anomalies and congenital communicating hydrocephalus (HYC2 2023). From the platform GeneCards, *MPDZ* mRNA express in normal expression pattern in human tissues from Serial Analysis of Gene Expression. Evidence shows that *MPDZ* is directly associated with Crumbs homolog 1 (Crb1), in which loss of Crb1 function leads to either recessively inherited retinitis pigmentosa or Leber congenital amaurosis in humans. The identification of human mutations suggests that *MPDZ* plays a role in human retinal disease, but the precise nature of this role remains to be determined (Ali et al. 2011). It has also been demonstrated that *MPDZ* enhances Notch signaling activity and that angiogenesis is coordinated by *VEGF* and Notch signaling (Tetzlaff et al. 2018). Inactivation of the *MPDZ* gene leads to impaired Notch signaling activity and increased blood vessel sprouting in cellular models (Tetzlaff et al. 2018), which may lead to increased vascularity in the DR. Since a recent study identified that *MPDZ* plays an essential role in the occurrence and maintenance of the macula (Zhang et al. 2022), a genetic correlation analysis of DR and macula can be performed in future studies.

It is worth mentioning that a suggestive locus rs17277029 in chromosome 4 with a *p* value of 6.01×10^−7^ in gene *FBXW7* had been found in primary GWAS. *FBXW7* was reported to be associated with DR, which in detail, in mouse experiments, silencing *FBXW7* enhanced angiogenesis and overexpressing *FBXW7* depressed angiogenesis in retinal tissue (Hu et al. 2020). Another study showed that the abundant mobilization of DNA damage repair mediated by *FBXW7* leads to the downregulation of poly-ADP-ribose-polymerase(PARP) expression and activity in both human endothelial cells and diabetic rat retinas, where RARP plays important roles in endothelial cell impairment in the pathogenesis of diabetic vascular diseases (Li et al. 2022). The GeneMANIA (https://genemania.org/) shows that *FBXW7* and *EYA2*, *NTNG1* had directly genetic interactions, with *MPDZ* had indirectly genetic interaction in Fig. 4. The remaining suggestive loci from other studies are not described in further detail here, but they hold certain value. For instance, primary GWAS(DoD>Q2) have shown that gene *LRP2* has been identified in relation to DR (Sheu et al. 2013), and *KCNJ3* from GWAS(+Urea) has been associated with the retina (Pattnaik et al. 2012). Moreover, Fig. 4 also shows in the gene linkage network, *RIMS2* serves as the intersection of *EYA2*, *MPDZ* and *NTNG1*, showing a certain association between them, which had reported potentially associated with DR. Data mining of human adult bulk and single-cell retinal transcriptional datasets revealed predominant expression in rod photoreceptors, and immunostaining demonstrated *RIMS2* localization in the human retinal outer plexiform layer, purkinje cells, and pancreatic islets (Mechaussier et al. 2020). A study shows that circular RNA is a potential target to control diabetic proliferative retinopathy (Shan et al. 2017), while circular *Rims2* (*circRims2*) is highly expressed and conserved in both the human and mouse brains and the high-throughput RNA-seq analysis reveals a high expression of *circRims2* in the retina (Sun et al. 2021). Furthermore, the current study finds that *circRims2* deficiency evokes retinal inflammation and activates the tumor necrosis factor signaling pathway (Sun et al. 2021), which is similar to that caused by inactivation of gene *MPDZ* (Tetzlaff et al. 2018). Although this is an alternative approach to using the identified loci to search for possible sources of mutations, there is no intuitive evidence that the mutation in the source gene *RIMS2* causes the development of DR in our GWAS studies.

**Fig. 4.**
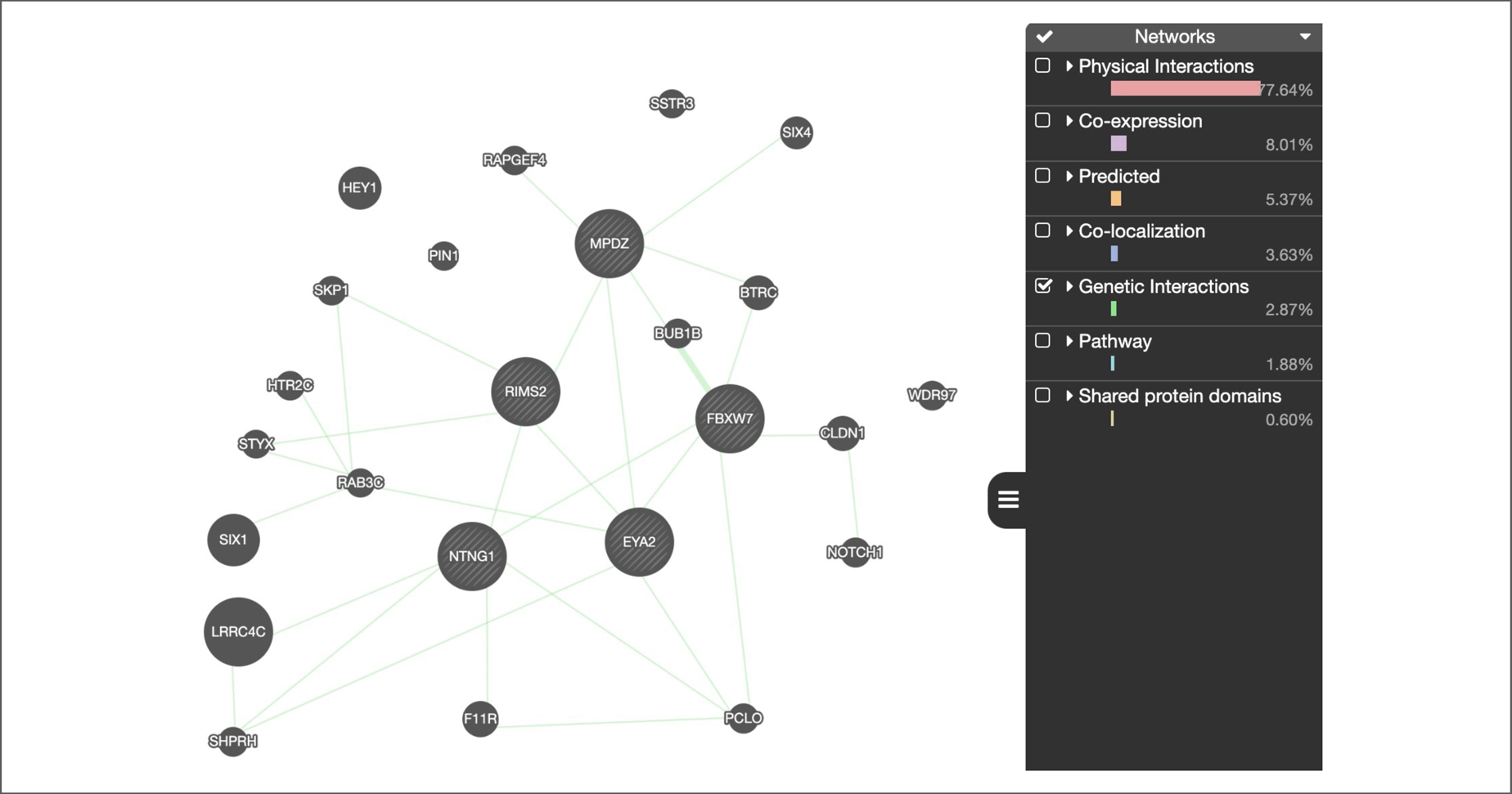
The genetic interaction network of the key genes from GeneMANIA

Our study, we defined T2D as the immediate use of insulin by the patient within one year of the detection of diabetes, a definition that was not present in UKB and carries some risk. We used multiple methods to define DR cases and controls datasets, so that doing different GWAS will not only improve data and study quality, but also help to discover new mechanisms of DR at the molecular level. For example, significant loci in primary GWAS showed more significant expression in GWAS (+HbA1c), suggesting that the inclusion of the covariate HbA1c helps to identify loci associated with DR, demonstrating the importance of this metric in studying DR. Our study, while providing initial insights, is not highly interpretive in designing different GWAS studies. The addition of covariates to the fastGWA-GLMM model is somewhat empirical, and the use of quartiles for dataset partitioning is somewhat subjective. This partitioning was necessitated in order to mine more information out of the limited dataset. Currently, most studies conduct GWASs using existing pipelines without considering the diversity of variable inputs into the model itself and the quality of model fitting. It is hoped that in the future, statistical knowledge can be utilized to explain the rationality, significance, and limitations of the fitted models more comprehensively. Furthermore, while we have highlighted the p-values of significant loci in four cohorts during the replication phase, it is essential to recognize that the statistical power of these results is relatively limited. In GoDARTS and GoSHARE, due to the limited number of T1D cases in the samples used, the disease background differs slightly from our study, which is entirely focused on T2D. In the study of Caucasian Australians cohort and Chinese cohort, the subject is PDR, which also differs from our research subject, DR. In FinnGen, the definition of controls applies to all participants, not just those restricted to having T2D.

Overall, this study suggests a role for the *EYA2* gene in retinal angiogenesis and proliferative retinopathy, with potential involvement of the spleen in retinal blood vessel repair. The *MPDZ* gene, possibly influencing Notch signaling, may promote retinal vascular sprouting in diabetic patients, while *FBXW7*’s overexpression depresses angiogenesis in retinal tissues. While limited evidence exists for other loci and neighboring genes, they offer reference points for future DR research. During the replication phase, no significant loci were replicated due to 1) the limited number of GWAS studies on DR, 2) the challenge of finding consistent subject definitions in GWAS studies targeting DR, and 3) the difficulty of accessing summary statistics for DR as many datasets remained unpublished. The replication cohorts that most closely matched the definition of our study were GoDARTS and GoSHARE, but direct replication was not achieved. This study highlights the *EYA2*, *MPDZ*, *NTNG1*, *CTAGE14P* and *MREGP1* genes, along with *FBXW7* and *LRP2*, for their higher association with DR in type 2 patients, helping to understand the mechanisms of DR and identify potential targets for DR treatment.

## Statements and Declarations

### Data availability

This research complies with all ethical standards and data privacy measures set by the UK Biobank. Detailed summary statistics data pertaining to Diabetic Retinopathy within the UK Biobank are retrievable from https://figshare.com/s/9d55aa774fb19307f7c8. Should there be any additional data pertinent to this study that are not presented within this paper or its additional files, the authors can provide such data upon reasonable request. Data is available upon publication.

### Authors’ Contributions

TC drafted the paper and performed GWAS analysis. QP, YT contributed to data formatting and correction. AR, CN, CP, TD, MH provided comments to the paper. WM organized the project and provided comments.

## Acknowledgment

The authors gratefully thank all the participants and professionals contributing to the UK Biobank. This research has been conducted using the UK Biobank Resource (50604).

## Consent to Publish

All authors have consent for publication.

## Ethics approval

This study was approved by the Ethics Committee of the University of Nottingham, Ningbo, China.

---

**Figure S1** The QQ plot of primary GWAS

**Figure S2** Manhattan plot of gene analysis in primary GWAS

**Figure S3** The regional plots of significant loci of five significant GWASs

**Figure S4** The QQ plots of additional GWASs

