## Supplementary figures and images for "Multiple genome-wide association studies of type 2 diabetes implicate several genes are associated with diabetic retinopathy based on UK Biobank"

### Figure S1.pdf

#### The QQ plot of primary GWAS

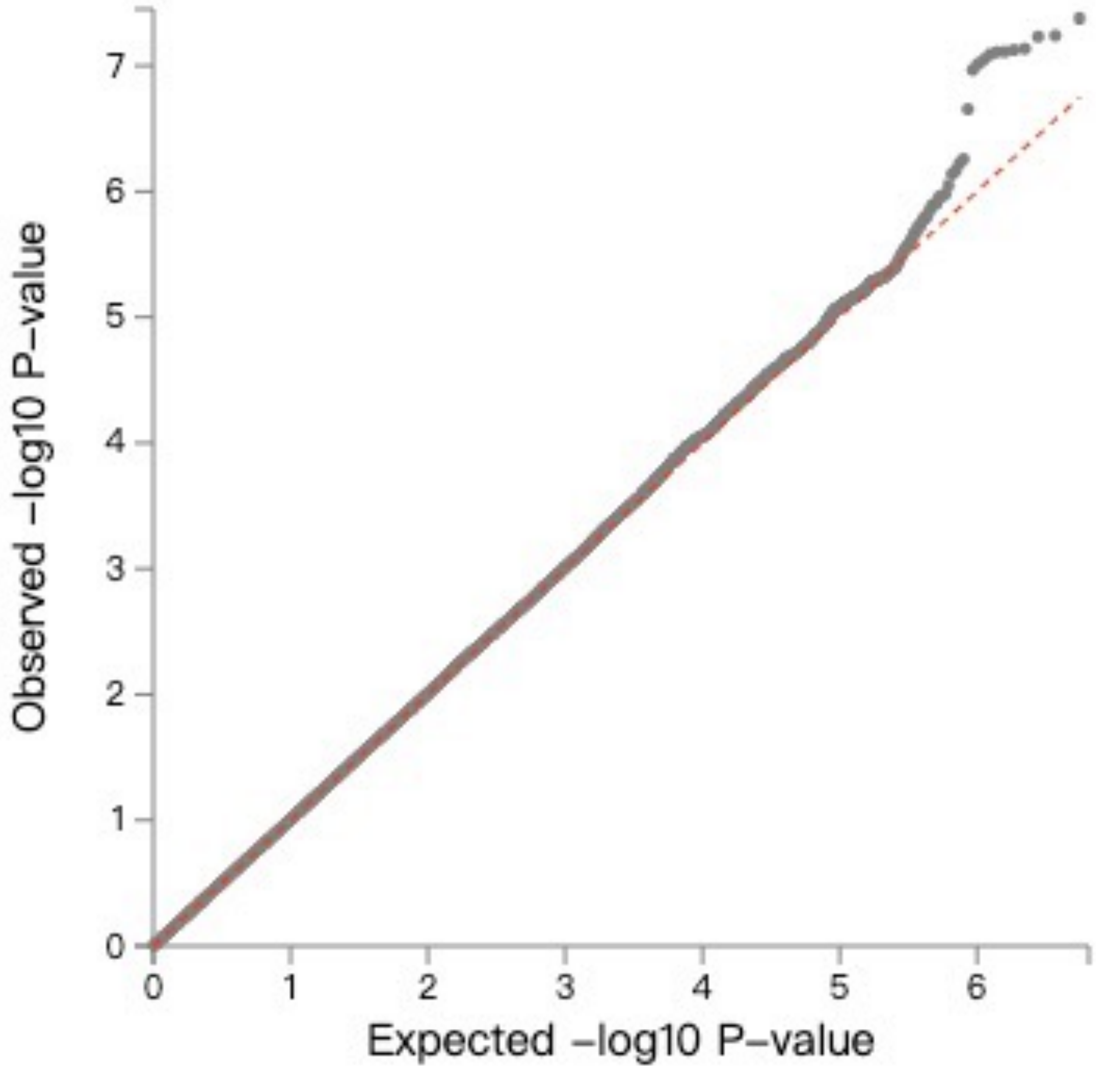

### Figure S2.pdf

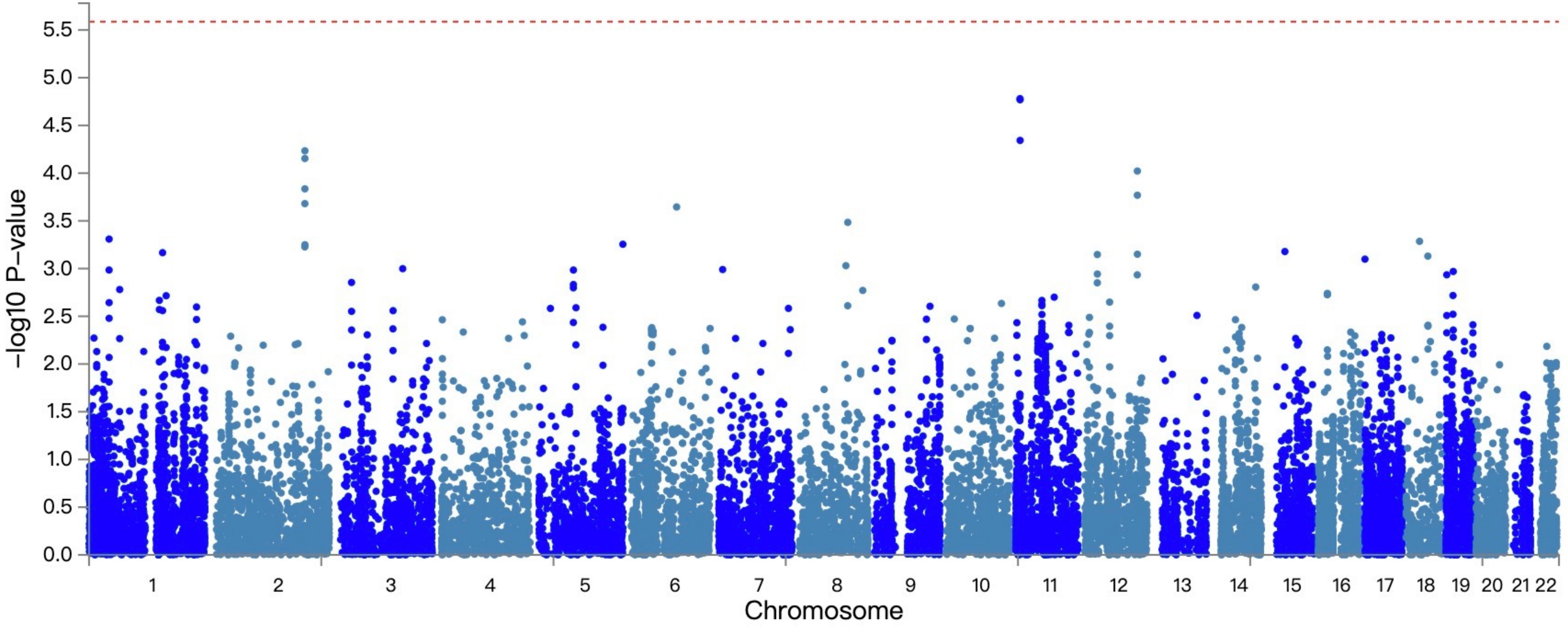

### Figure S3.pdf

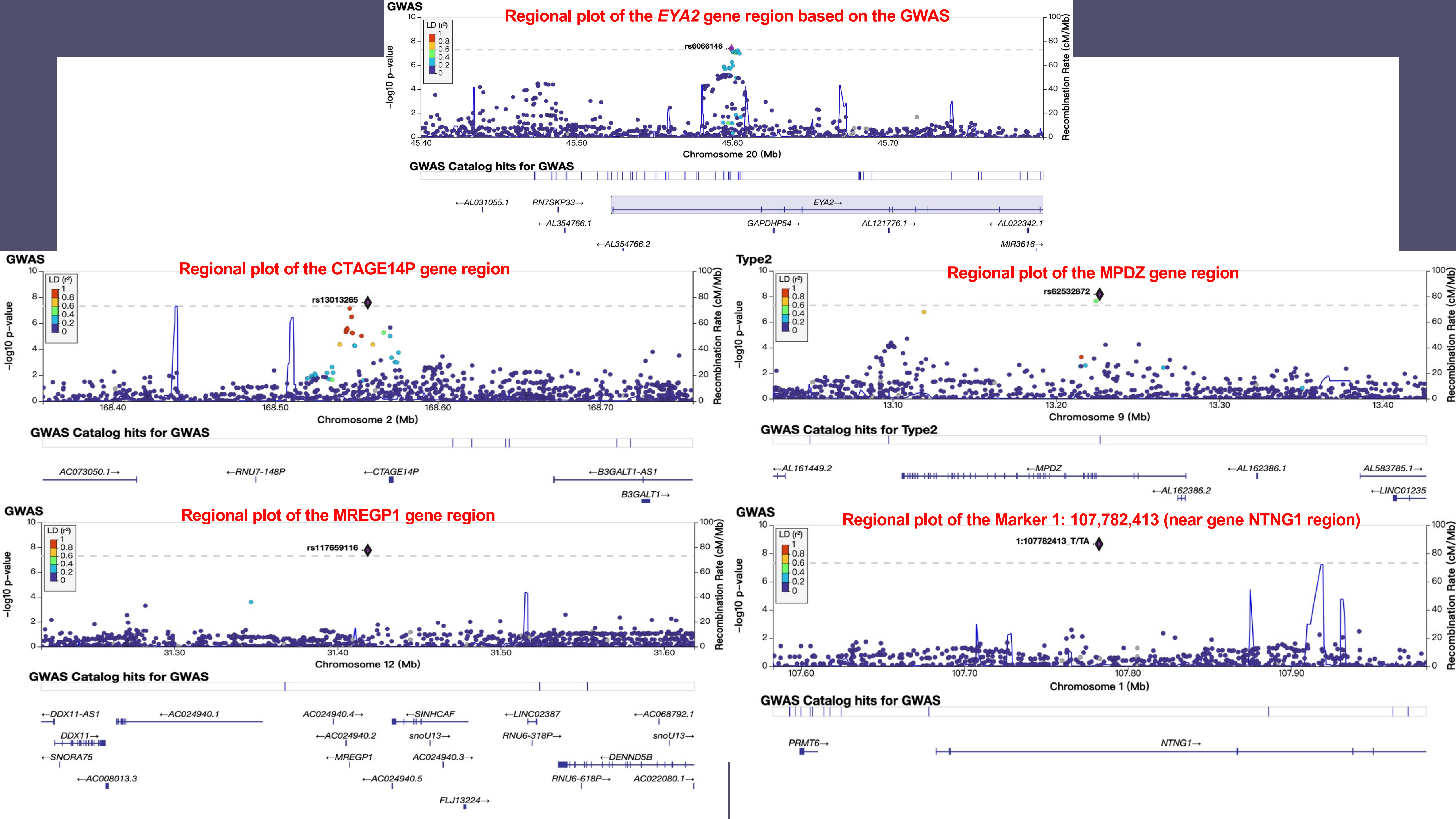

### Figure S4.pdf

QQ plot of GWAS(+HbA1c)

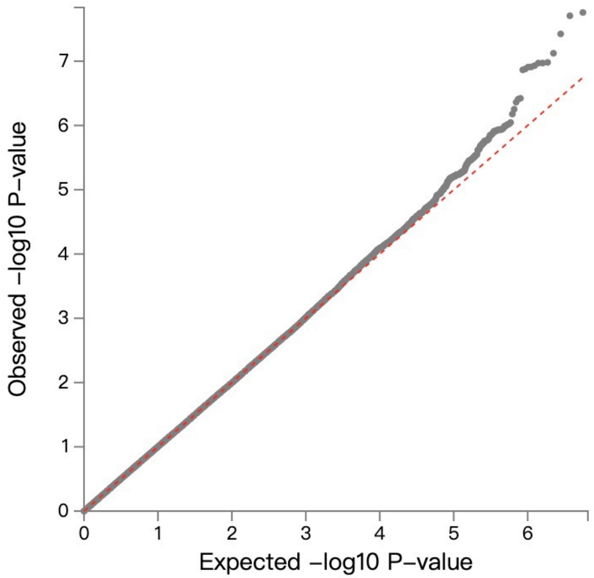


QQ plot of primary GWAS(DoD>Q2)

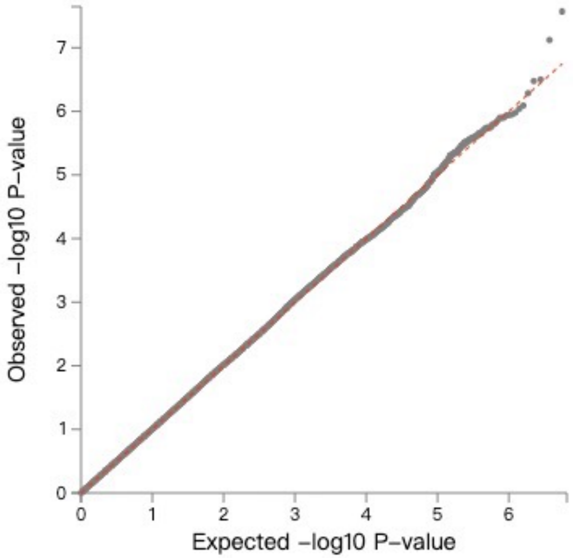


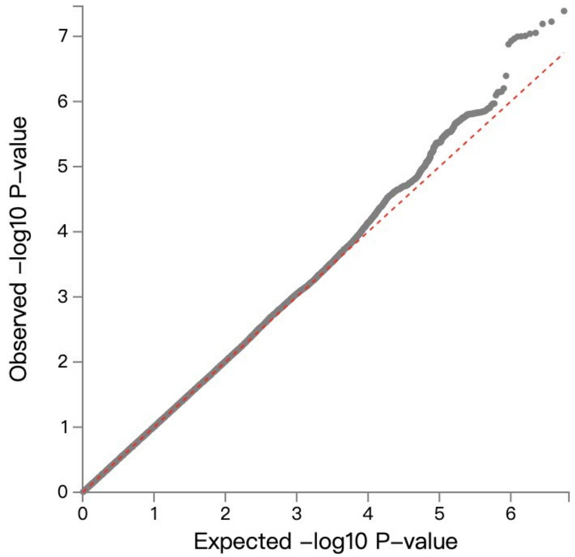


QQ plot of GWAS(+Urea)

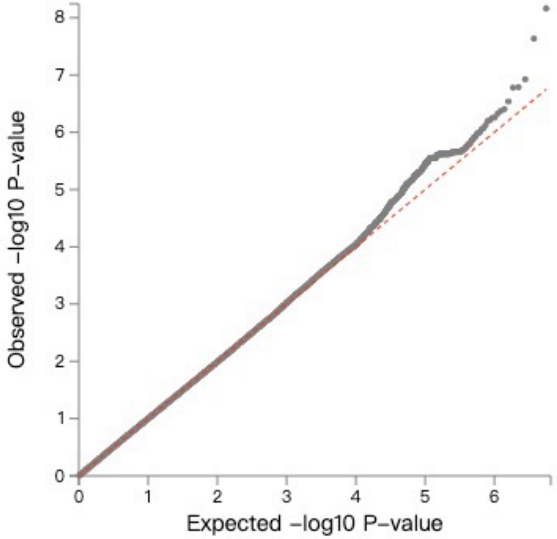


QQ plot of GWAS(HbA1c>Q3)
